# Reduced serum carboxymethyllysine is independently associated with Alzheimer’s disease

**DOI:** 10.64898/2026.11.08.26367562

**Authors:** Yihong Li, Mei Yang, Xin Feng, Guosheng Gao, Huadong Fan

## Abstract

**INTRODUCTION:** Advanced glycation end products accumulate in Alzheimer’s disease (AD) brain, but the clinical significance of circulating N^ε^-(carboxymethyl)lysine (CML) remains uncertain. We evaluated serum CML as a complementary peripheral biomarker of AD.

**METHODS:** This observational case–control study enrolled 101 patients with clinically diagnosed AD and 91 healthy controls. Serum CML was quantified in 191 participants. Multivariable logistic regression and receiver operating characteristic analyses assessed association and discrimination. A randomly selected subcohort underwent ultrasensitive measurement of amyloid-, tau-, glial-, and neuroaxonal-related biomarkers.

**RESULTS:** Serum CML was lower in AD than controls (median 15.26 versus 23.72 ng/mL; P<0.001). In a complete-case model (n=178), each 1-ng/mL increase was associated with lower odds of AD (adjusted odds ratio 0.727, 95% confidence interval 0.656–0.804). CML alone yielded an area under the curve (AUC) of 0.883. Five-fold cross-validation yielded AUCs of 0.727 for a clinical model and 0.911 after adding CML. In the biomarker subcohort, AD showed lower Aβ42/Aβ40 and higher p-tau217; exploratory CML plus p-tau217 discrimination reached an AUC of 0.966 (n=60).

**DISCUSSION:** Reduced serum CML was independently associated with clinically diagnosed AD and may reflect a systemic glycoxidative dimension complementary to canonical biomarkers. Its biological origins and clinical value require external, longitudinal, and biomarker-defined validation.

## Introduction

Alzheimer’s disease (AD), the leading cause of dementia, is biologically characterized by amyloid-β (Aβ) deposition and tau neurofibrillary pathology [1, 2]. Contemporary diagnostic frameworks increasingly incorporate fluid and imaging biomarkers to characterize underlying AD pathology [3, 4]. Blood-based biomarkers, particularly phosphorylated tau217 (p-tau217), have demonstrated strong diagnostic performance in appropriately selected clinical populations [5, 6]. Nevertheless, AD involves broader biological disturbances, including oxidative stress, neuroinflammation, mitochondrial dysfunction, and systemic metabolic dysregulation, which are not fully captured by amyloid- and tau-centered biomarkers [7]. Identifying complementary peripheral biomarkers may therefore provide additional insights into disease-associated systemic processes.

Advanced glycation end products (AGEs) comprise chemically heterogeneous modifications generated through nonenzymatic glycation, glycoxidation, and reactive carbonyl chemistry [8]. Their circulating abundance reflects a dynamic balance among endogenous formation, dietary exposure, protein turnover, tissue retention, and renal clearance [9, 10]. N^ε^-(carboxymethyl)lysine (CML), a prevalent and extensively studied AGE modification, is widely regarded as an indicator of glycoxidative stress [11]. However, circulating CML concentrations are influenced by aging, oxidative injury, metabolic status, dietary intake, and kidney function [12, 13]. These multiple determinants complicate interpretation of circulating CML as a disease-associated biomarker and highlight the importance of accounting for potential confounding factors.

Neuropathological investigations have demonstrated substantial accumulation of CML in the aging and AD brain, although its distribution is anatomically and cellularly heterogeneous. Early studies identified CML immunoreactivity within neuronal perikarya and extraneuronal deposits, frequently associated with intracellular lipofuscin [14]. Subsequent investigations revealed CML deposition in neurons, astrocytes, and microglia, with particularly pronounced glial accumulation in AD [15, 16]. Regionally, CML immunoreactivity is prominent in the cerebral cortex and hippocampus but relatively limited in several subcortical structures. Within the hippocampus, CA3 and CA4 exhibit greater CML immunoreactivity than CA1 and CA2, while extraneuronal deposits are frequently associated with astroglial structures [15–17]. Furthermore, CML co-localizes with malondialdehyde, a lipid-peroxidation-associated product, suggesting an intersection between glycoxidative modification and oxidative lipid injury [18].

Importantly, cerebral CML deposition does not necessarily parallel conventional AD neuropathology. Neurons containing abundant neurofibrillary tangles may exhibit minimal CML immunoreactivity, whereas CML-rich neurons can occur in regions with comparatively limited tau pathology or neuronal loss. An inverse relationship between regional CML deposition and Braak stage has also been reported [15, 17]. At the molecular level, site-specific CML modification of Aβ42 at Lys16 or Lys28 reduces fibrillar aggregation while increasing cytotoxicity, potentially favoring persistent toxic oligomeric intermediates rather than enhanced reactive oxygen species generation [19]. These observations suggest that the biological consequences of CML depend on its molecular targets, modification sites, and cellular localization.

The relationship between cerebral and circulating CML remains incompletely understood. Experimental evidence implicates age-associated intestinal barrier dysfunction and microbiota-dependent CML exposure in cerebral accumulation, microglial oxidative stress, and mitochondrial impairment [20]. Clinical investigations have reported increased cerebrospinal fluid CML without a corresponding increase in serum concentrations [21]. Other studies identified elevated CML in albumin-free serum proteins during early AD but reduced extracellular-vesicle-associated CML in moderate disease [22], whereas increased plasma CML and sex-dependent variation have also been described [23]. Differences in biological matrices, molecular fractions, analytical methodologies, disease stages, and metabolic or renal characteristics may contribute to these apparently conflicting findings. Consequently, the direction and independent clinical significance of circulating CML alterations in AD remain unresolved.

In this observational case–control study, we measured serum CML in 101 clinically diagnosed AD patients and 91 healthy controls and examined its association with AD after accounting for demographic, renal, glycemic, and metabolic factors. We also evaluated its discriminatory contribution beyond routine clinical variables. In a randomly selected subcohort, amyloid biomarkers (Aβ42, Aβ40, and Aβ42/Aβ40) served as principal biological comparators, p-tau217 as the primary exploratory tau marker, and p-tau181, GFAP, and NfL as supportive measures. We hypothesized that circulating CML captures a systemic glycoxidative/metabolic dimension complementary to canonical AD pathology.

## Methods

### 1.1 Study design and participants

This observational case–control biomarker study included 101 patients with clinically diagnosed AD and 91 healthy controls (HC) (n=192). Clinical and routine laboratory information was obtained from hospital examination records. Eligible participants were aged 60 years or older and included patients with clinically diagnosed AD and cognitively HC who voluntarily participated and provided written informed consent. Participants with non-AD cognitive disorders or other major neurological diseases, including stroke or vascular dementia, Parkinson’s disease, and frontotemporal dementia, were excluded, as were individuals with documented CMV, EBV, HIV, HCV, or HBV infection according to the approved study protocol. Peripheral venous blood (5 mL) was collected and serum was separated for analysis. Serum CML was available in 191 participants (101 AD and 90 HC), constituting the primary CML cohort for group comparison, multivariable logistic regression, and ROC analysis. A randomly selected biomarker subcohort comprised 31 AD participants and 29 HC (n=60) for ultrasensitive single-molecule array (Simoa) testing of Aβ42, Aβ40, p-tau181, p-tau217, GFAP, and NfL. Exploratory subcohort analyses included biomarker group comparisons and combined ROC analyses evaluating CML with Aβ42/Aβ40 or p-tau217. Figure 1 summarizes participant flow, data collection, and the primary and exploratory analytical pathways.

**Figure 1.**
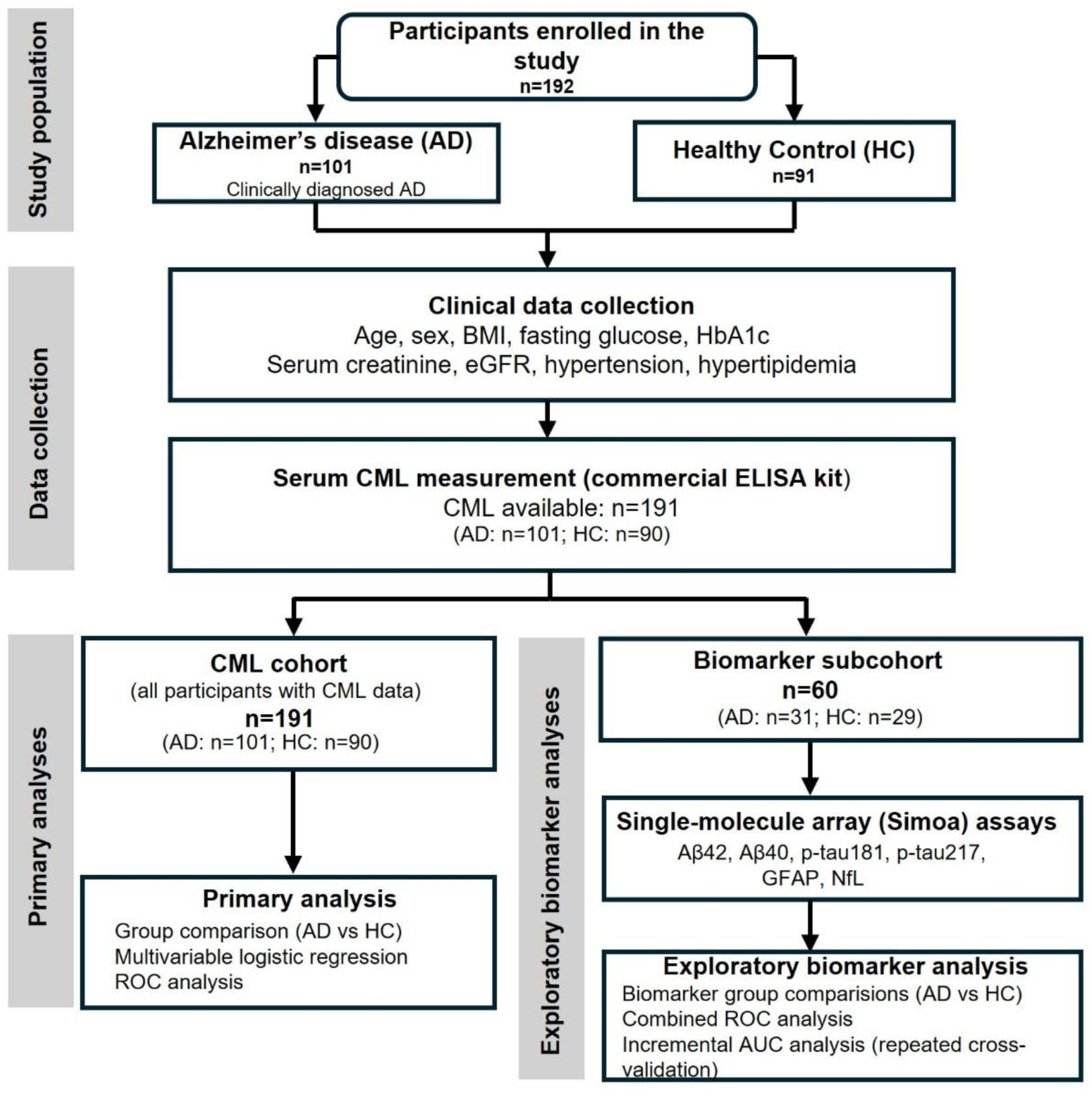
Study population and analytical workflow. A total of 192 participants were enrolled, including 101 patients with clinically diagnosed Alzheimer’s disease (AD) and 91 healthy controls (HC). Clinical data collection included age, sex, body mass index (BMI), fasting glucose, glycated hemoglobin (HbA1c), serum creatinine, estimated glomerular filtration rate (eGFR), hypertension, and hyperlipidemia. Serum CML was measured using a commercial ELISA kit and was available for 191 participants (101 AD and 90 HC), who constituted the primary CML cohort for group comparison, multivariable logistic regression, and ROC analysis. A randomly selected biomarker subcohort comprised 60 participants (31 AD and 29 HC) and underwent single-molecule array (Simoa) assays for Aβ42, Aβ40, p-tau181, p-tau217, GFAP, and NfL. Exploratory biomarker analyses included AD–HC group comparisons and combined ROC analyses assessing CML together with Aβ42/Aβ40 or p-tau217; incremental model performance was evaluated within the cross-validation framework described in the Statistical analysis section.

### 1.2 Ethics statement

The study was reviewed and approved by the Ethics Committee of Ningbo Institute of Life and Health Industry, University of Chinese Academy of Sciences (Approval No. SL-NBYJY-KY-2023-010-01) and the Ethics Committee of Biomedical Research Involving Humans, Ningbo University Affiliated Kangning Hospital (Approval No. NBKNYY-2024-LC-03). All procedures involving human participants were conducted in accordance with the Declaration of Helsinki and applicable institutional requirements. Written informed consent was obtained from all participants before enrollment and blood collection. Participants were informed of the study procedures, the voluntary nature of participation, their right to withdraw consent at any time without affecting medical care or other rights, and the planned use of their clinical data and biospecimens for research. Samples and study records were coded with study identifiers rather than participant names, and personally identifiable information was kept confidential and accessible only to authorized study personnel and, where required, regulatory or ethics authorities. Research data were retained by Ningbo Institute of Life and Health Industry, University of Chinese Academy of Sciences, in accordance with the approved protocol.

### 1.3 Clinical and laboratory variables

Demographic and routine clinical variables were retrieved from hospital records, including age, sex, height, weight, body mass index (BMI), fasting blood glucose, glycated hemoglobin (HbA1c), serum creatinine, estimated glomerular filtration rate (eGFR), hypertension, and hyperlipidemia. Missingness varied across routinely collected variables.Therefore, available-case denominators are reported for descriptive analyses and complete-case samples are specified for multivariable models.

### 1.4 Measurement of serum CML

Serum CML concentrations were quantified using a commercially available Carboxymethyllysine (CML) ELISA Kit (abx575945; Abbexa, UK) according to the manufacturer’s instructions. CML measurements were available for all 101 AD samples and 90 of 91 HC samples. Because CML exists both as a free lysine-derived adduct and covalently attached to circulating proteins, we additionally characterized the molecular species detected by the commercial kit and by the in-house assay described below.

An in-house indirect competitive enzyme-linked immunosorbent assay (ELISA) was established to provide an independent assessment of serum CML. The coating antigen was prepared by reacting bovine serum albumin (BSA; Absin, Shanghai, China) with glyoxal (Aladdin, Shanghai, China) in 0.2 M phosphate buffer (pH 8.0; Macklin, Shanghai, China) at 37 °C for 4 days in the dark. The reaction mixture was subsequently dialyzed against purified water for 24 h using a dialysis membrane with a molecular-weight cutoff of 8–14 kDa. For the competitive immunoassay, microplates were coated with the prepared CML-modified antigen and incubated with anti-CML primary antibody (ab125145; Abcam, Cambridge, UK) in the presence of CML standards or serum samples. Bound antibodies were detected using an anti-mouse IgG secondary antibody (Cell Signaling Technology, Danvers, MA, USA), followed by color development with tetramethylbenzidine (TMB). Absorbance was measured at 450 nm using a UV–VIS microplate spectrophotometer (SpectraMax; Molecular Devices, CA, USA).

The analytical target profiles of the two assays were further examined using free CML and CML-modified BSA (CML-BSA) standards (Supplementary Figure 3). In the in-house competitive assay, increasing concentrations of either free CML or CML-BSA produced concentration-dependent decreases in OD450, demonstrating recognition of both free CML and protein-bound CML epitopes (Supplementary Figure 3A, B). In contrast, when CML-BSA was tested with the commercial CML kit, OD450 remained essentially unchanged across the tested CML-BSA concentration range, indicating little detectable response to protein-bound CML under these assay conditions (Supplementary Figure 3C). Accordingly, the commercial kit predominantly reflects a free-CML-sensitive serum pool, whereas the in-house assay detects a broader immunoreactive pool comprising free and protein-bound CML. Serum CML measurements obtained using the two assays were compared in 60 paired serum samples (30 AD and 30 HC), and their association was evaluated using Spearman rank correlation analysis. Because the assays quantify different molecular pools and use different calibration systems, their absolute concentrations and reported units are not directly interchangeable. The cross-assay comparison was therefore intended to assess concordance in relative biological variation rather than numerical equivalence (Supplementary Figure 2).

### 1.5 Single-molecule immunoassay biomarker subcohort

Serum samples were randomly selected from 31 patients with AD and 29 HC for ultrasensitive single-molecule biomarker analysis. Serum concentrations of AD-related biomarkers, including phosphorylated tau at threonine 181 (p-tau181), phosphorylated tau at threonine 217 (p-tau217), glial fibrillary acidic protein (GFAP), neurofilament light chain (NfL), amyloid-β42 (Aβ42), and amyloid-β40 (Aβ40), were quantified using an ultrasensitive single-molecule array (Simoa) immunoassay platform (BioChip Reader, Iomics Bioscience Inc., Beijing, China). Serum samples were stored at −80 °C until analysis, and biomarker measurements were performed according to the manufacturer’s instructions. For quantitative summaries, assay results reported below the lower limit of quantification were assigned the corresponding limit values. The Aβ42/Aβ40 ratio was calculated from the quantitative Aβ42 and Aβ40 values. Amyloid measures were prespecified as the principal biological comparator because they directly complement the CML-focused hypothesis. p-tau217 was retained as the principal exploratory tau marker because of its high performance for AD pathology. p-tau181, GFAP, and NfL were analyzed as supportive exploratory measures and were not used to redefine the primary study endpoint. Exploratory combined analyses paired CML with Aβ42/Aβ40 and, separately, with p-tau217 to assess potential complementarity across systemic glycoxidative, amyloid-related, and tau-related signals. Analyte-specific sample sizes are reported because the final analysis sets differed slightly across biomarkers. Vendor-supplied reference intervals that were not validated for serum were not used to classify participants as biomarker-positive or biomarker-negative.

### 1.6 Statistical analysis

Continuous variables were summarized as median and interquartile range (IQR) and compared using two-sided Mann–Whitney U tests. Categorical variables were summarized as n (%) and compared using χ² tests or Fisher’s exact test when appropriate. The association between CML and AD was evaluated by logistic regression. Sequential sensitivity models expressed CML per 1-SD increase and adjusted for age and sex, then eGFR, and finally fasting glucose, HbA1c, BMI, hypertension, and hyperlipidemia. A parsimonious multivariable model was additionally fitted using CML in its original unit (ng/mL) together with age, sex, eGFR, and HbA1c to provide directly interpretable adjusted effects and to support the clinical-model ROC analysis. Complete-case analysis was used within each model. In the single-molecule subcohort, biomarker distributions were summarized as median (IQR) and AD–HC differences were assessed using two-sided Mann–Whitney U tests, with analyte-specific sample sizes reported in Supplementary Table 1 and Figure 5. To examine whether renal function could account for the CML difference, Spearman rank correlations between serum CML and eGFR were calculated separately in AD and healthy controls (Supplementary Figure 1).

Receiver operating characteristic (ROC) curves were used to assess discrimination. For CML alone in the full CML cohort (n=191), the AUC and 95% confidence interval (CI) were estimated by bootstrap resampling. For comparison of the clinical model (age, sex, eGFR, and HbA1c) with the clinical model plus CML, analyses were restricted to 178 participants with complete data (101 AD and 77 HC). ROC curves for these fitted models were generated from out-of-fold predictions using stratified five-fold cross-validation; the corresponding AUCs were 0.727 and 0.911, respectively. In the single-molecule subcohort, two exploratory ROC analyses were performed using the complete cases available for each biomarker pairing: CML, Aβ42/Aβ40, and a CML + Aβ42/Aβ40 model (n=59; 30 AD and 29 controls), and CML, p-tau217, and a CML + p-tau217 model (n=60; 31 AD and 29 controls). Model performance in these exploratory combined analyses was evaluated using stratified five-fold cross-validation. Because the CML AUC was calculated within the complete-case sample for each biomarker pairing, its value differed slightly between the two subcohort analyses. Spearman’s rank correlation was used to assess the association between serum CML and eGFR separately in AD and HC. Two-sided P<0.05 was considered statistically significant.

## Results

### 1.1 Study population and clinical characteristics

To characterize the study population and identify demographic, renal, and metabolic factors that might influence circulating CML concentrations or confound its association with AD, we first compared the clinical characteristics of the AD and healthy control groups. A total of 192 participants were included, comprising 101 patients with AD and 91 HC (Figure 1).

The AD group was older than the control group [median 74.0 years (IQR 69.0–77.0) versus 68.0 years (64.0–71.5), P<0.001], whereas sex distribution did not differ significantly (male: 36.6% versus 47.3%, P=0.179). BMI, fasting glucose, HbA1c, and serum creatinine were also comparable between groups. eGFR was modestly lower in AD [91.98 (86.47–95.03) versus 95.38 (85.94–100.68) mL/min/1.73 m²; P=0.006], whereas hyperlipidemia was less frequent in AD (P=0.003). Given their potential relevance to circulating CML homeostasis, these variables were considered in subsequent multivariable analyses where appropriate (Table 1).

**Table 1.**
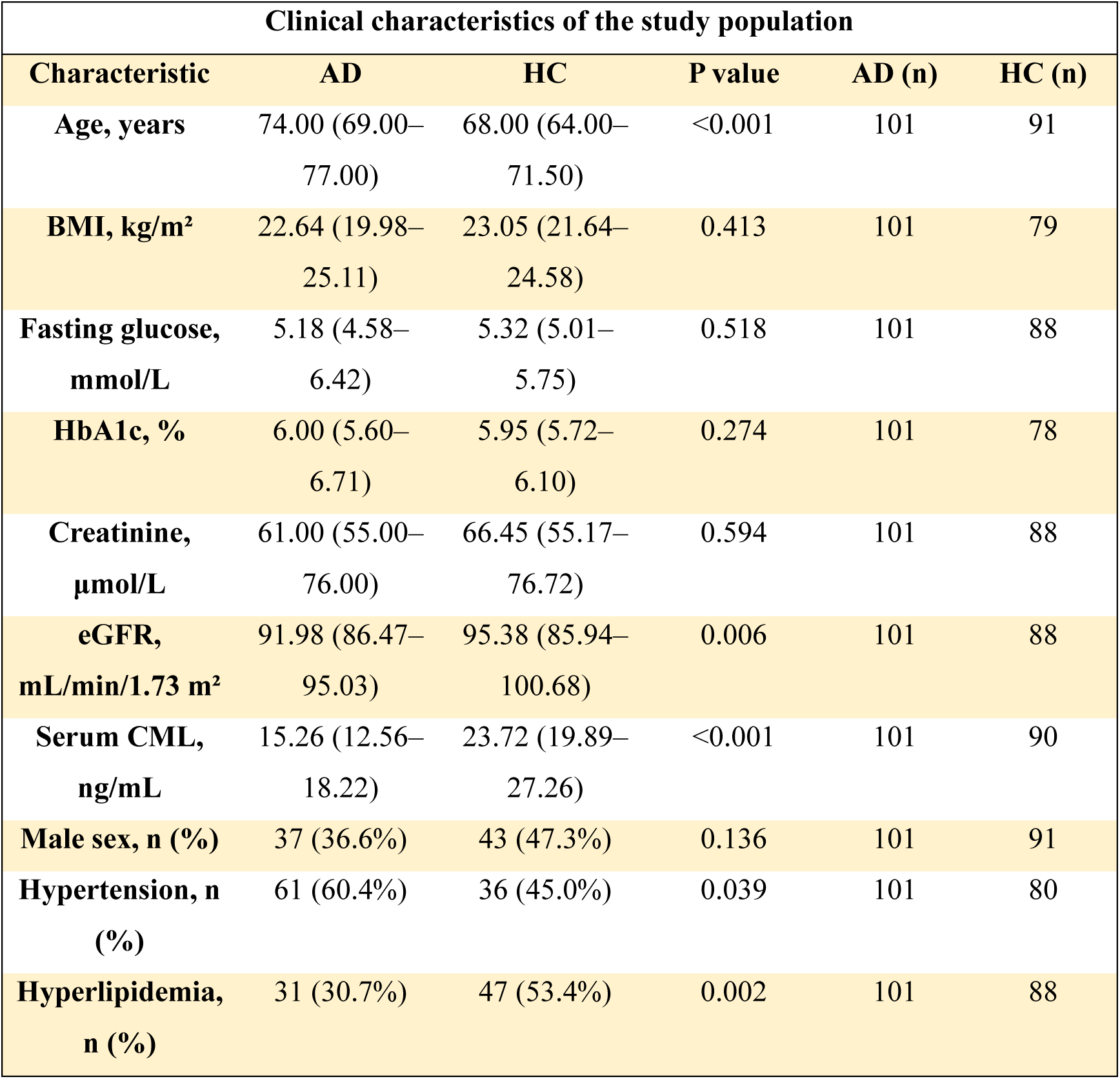
Clinical characteristics of the study population. Continuous variables are median (IQR) and were compared using two-sided Mann–Whitney U tests. Categorical variables are n (%) and were compared using χ² tests (or Fisher’s exact test where appropriate). Available-case denominators are reported because some routine variables were missing. AD, Alzheimer’s disease; BMI, body mass index; CML, N^ε^-(carboxymethyl)lysine; eGFR, estimated glomerular filtration rate; HbA1c, glycated hemoglobin.

### 1.2 Serum CML is markedly reduced in Alzheimer’s disease

Having characterized the clinical profiles of the two groups, we next examined whether circulating CML differed between patients with AD and HC. CML measurements were available in 191 participants (101 AD and 90 controls). Serum CML measured with the commercial kit was markedly lower in AD than in controls [15.26 (12.56–18.22) versus 23.72 (19.89–27.26) ng/mL; P<0.001] (Figure 2; Table 1). The distributions showed substantial separation between groups despite some overlap, establishing a pronounced unadjusted difference in the commercial-kit CML signal associated with AD status. To independently assess the direction of this finding, an in-house competitive ELISA was applied to a subset of 60 serum samples (30 AD and 30 HC) and similarly showed lower CML in AD (Supplementary Figure 2A). Measurements from the two assays were moderately positively correlated (Spearman ρ=0.572, P<0.001; Supplementary Figure 2B), despite a marked difference in absolute concentration scale. Analytical characterization clarified this discrepancy: the in-house assay responded to both free CML and CML-BSA, whereas the commercial kit showed little detectable response to CML-BSA across the tested concentration range (Supplementary Figure 3). Thus, the two assays capture overlapping but non-identical CML pools: the in-house assay reflects a broader free-plus-protein-bound immunoreactive CML signal, while the commercial kit predominantly reflects a free-CML-sensitive pool. Their agreement in the direction of the AD–HC difference and their positive rank correlation therefore support biological concordance without implying equivalence of absolute concentration or units.

**Figure 2.**
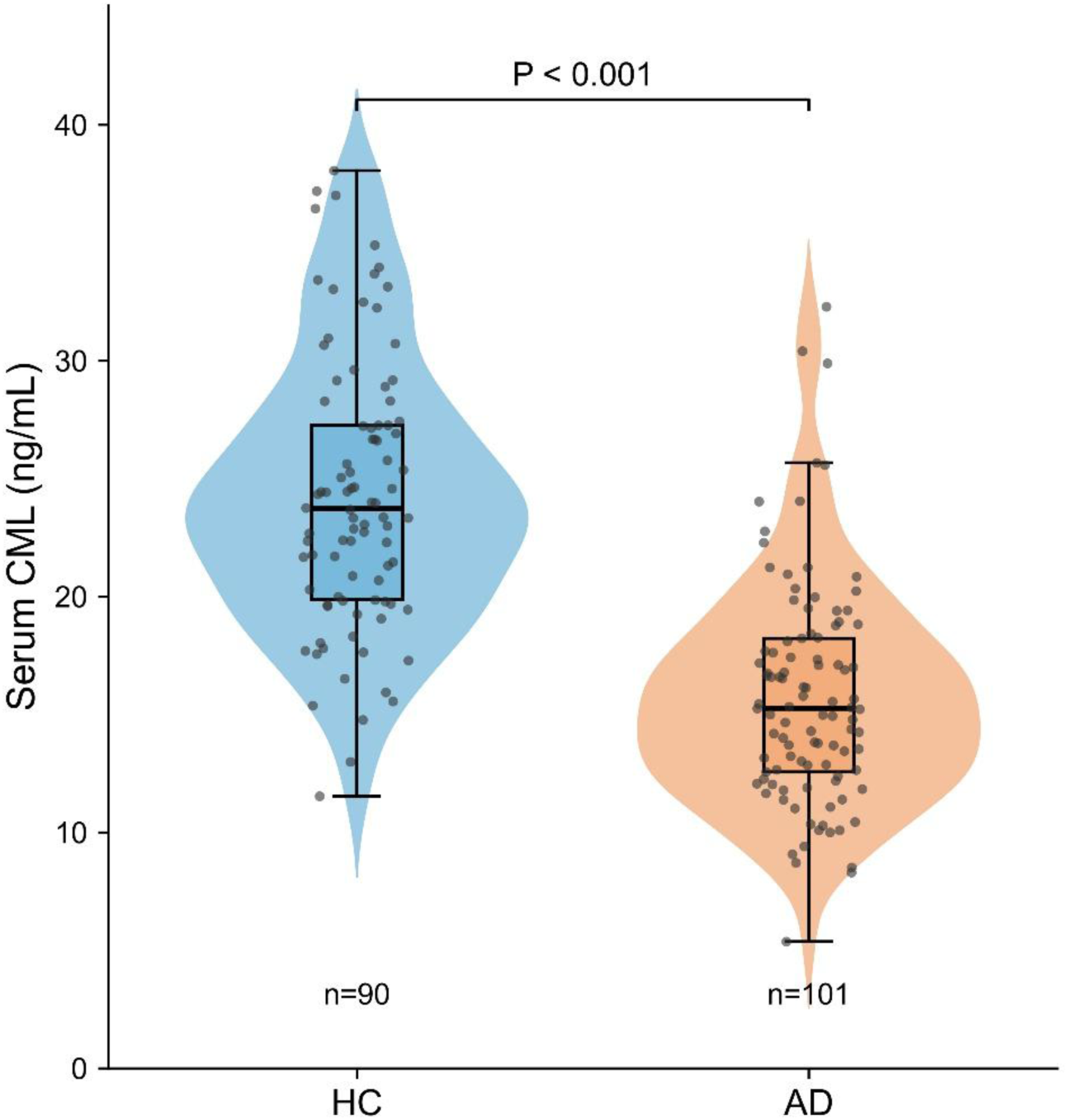
Serum CML is markedly reduced in Alzheimer’s disease. Violin plots showing the distribution of serum CML concentrations in patients with Alzheimer’s disease (AD, n=101) and healthy controls (HC, n=90). Violin widths represent the estimated data density and individual dots indicate individual participants. The embedded box plots indicate the median and interquartile range (IQR), with whiskers extending to the most extreme observations within 1.5×IQR. Statistical significance was assessed using a two-sided Mann–Whitney U test.

### 1.3 The inverse CML–AD association persists after multivariable adjustment

Because circulating CML can be influenced by age, renal function, glycemic status, and other metabolic factors, we next tested whether the observed CML–AD association persisted after accounting for these potential confounders. Across sequential logistic regression models, higher serum CML remained strongly associated with lower odds of AD. The OR per 1-SD higher CML was 0.123 (95% CI 0.068–0.222) in the unadjusted model, 0.140 (0.076–0.256) after adjustment for age and sex, 0.139 (0.076–0.256) after additional adjustment for eGFR, and 0.120 (0.057–0.251) in the fully adjusted complete-case model that additionally included fasting glucose, HbA1c, BMI, hypertension, and hyperlipidemia (all P<0.001; Table 2).

**Table 2.** Sequential association of serum CML with Alzheimer’s disease. ORs represent the change in odds of AD per 1-SD higher serum CML. Model 1 adjusts for age and sex; Model 2 additionally adjusts for eGFR; Model 3 additionally adjusts for fasting glucose, HbA1c, BMI, hypertension, and hyperlipidemia. Complete-case analysis was used within each model. OR, odds ratio; CI, confidence interval.

| **Model** | **Adjustment** | **n** | **OR per 1-SD CML** | **95% CI** | **P value** |
|---|---|---|---|---|---|
| **Unadjusted** | None | 191 | 0.123 | 0.068–0.222 | <0.001 |
| **Model 1** | Age, sex | 191 | 0.140 | 0.076–0.256 | <0.001 |
| **Model 2** | Age, sex, eGFR | 188 | 0.139 | 0.076–0.256 | <0.001 |
| **Model 3** | Age, sex, eGFR, glucose, HbA1c, BMI, hypertension, hyperlipidemia | 169 | 0.120 | 0.057–0.251 | <0.001 |

To provide an effect estimate directly interpretable in the original CML concentration scale and aligned with the subsequent clinical-model analysis, we additionally fitted a parsimonious complete-case model including CML, age, sex, eGFR, and HbA1c. Among 178 participants with complete data (101 AD and 77 healthy controls), each 1-ng/mL higher CML was associated with 27.3% lower adjusted odds of AD (OR 0.727, 95% CI 0.656–0.804; P<0.001), whereas age, sex, eGFR, and HbA1c were not independently significant (Figure 3). Thus, the strong inverse association between serum CML and AD remained evident across models with progressively broader adjustment for major measured demographic, renal, glycemic, and metabolic factors. Because renal function is an important determinant of circulating CML, we also examined the relationship between serum CML and eGFR within each diagnostic group. Among participants with paired measurements, CML was not significantly correlated with eGFR in either AD (n=101; Spearman ρ=0.012, P=0.904) or HC (n=87; ρ=0.202, P=0.060) (Supplementary Figure 1). Together with the multivariable models, these findings argue against the between-group CML difference being primarily attributable to measured renal function.

**Figure 3.**
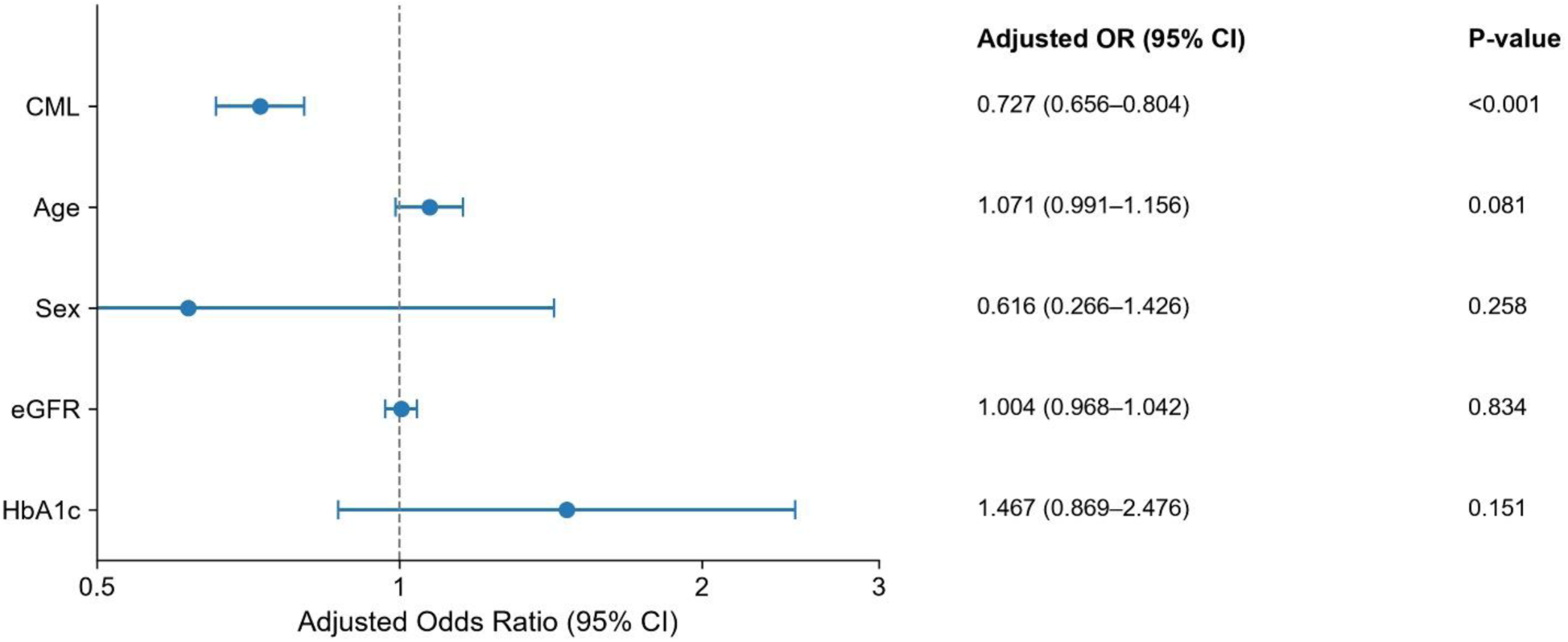
Association of serum CML with Alzheimer’s disease after multivariable adjustment. Forest plot showing adjusted odds ratios (ORs) and 95% confidence intervals (CIs) from a multivariable logistic regression model including serum CML, age, sex, estimated glomerular filtration rate (eGFR), and glycated hemoglobin (HbA1c). The analysis included 178 participants (101 AD and 77 HC) with complete data. ORs represent the change in odds of AD per 1-ng/mL increase in CML, 1-year increase in age, 1-mL/min/1.73 m² increase in eGFR, and 1-percentage-point increase in HbA1c. For sex, the OR represents males relative to females (reference group). Points indicate adjusted ORs, and horizontal bars represent 95% CIs. The dashed vertical line indicates an OR of 1.

### 1.4 CML provides substantial discrimination and adds information to a clinical model

Given the robust independent association between serum CML and AD, we next assessed its ability to discriminate patients with AD from HC and whether it provided information beyond routinely available clinical variables. In the full CML cohort (n=191), CML alone yielded an AUC of 0.883 (95% CI 0.830–0.925; Figure 4A). We then compared a clinical model incorporating age, sex, eGFR, and HbA1c with the same model additionally including CML. This analysis included 178 participants (101 AD and 77 healthy controls) with complete data for all model variables. Using out-of-fold predictions from stratified five-fold cross-validation, the clinical model yielded an AUC of 0.727 (95% CI 0.649–0.799), whereas addition of CML increased the AUC to 0.911 (95% CI 0.861– 0.953; Figure 4B). These cross-validated results indicate that CML contributed substantial discriminatory information beyond the selected demographic, renal, and glycemic variables in this cohort, while the generalizability of this improvement requires independent external validation.

**Figure 4.**
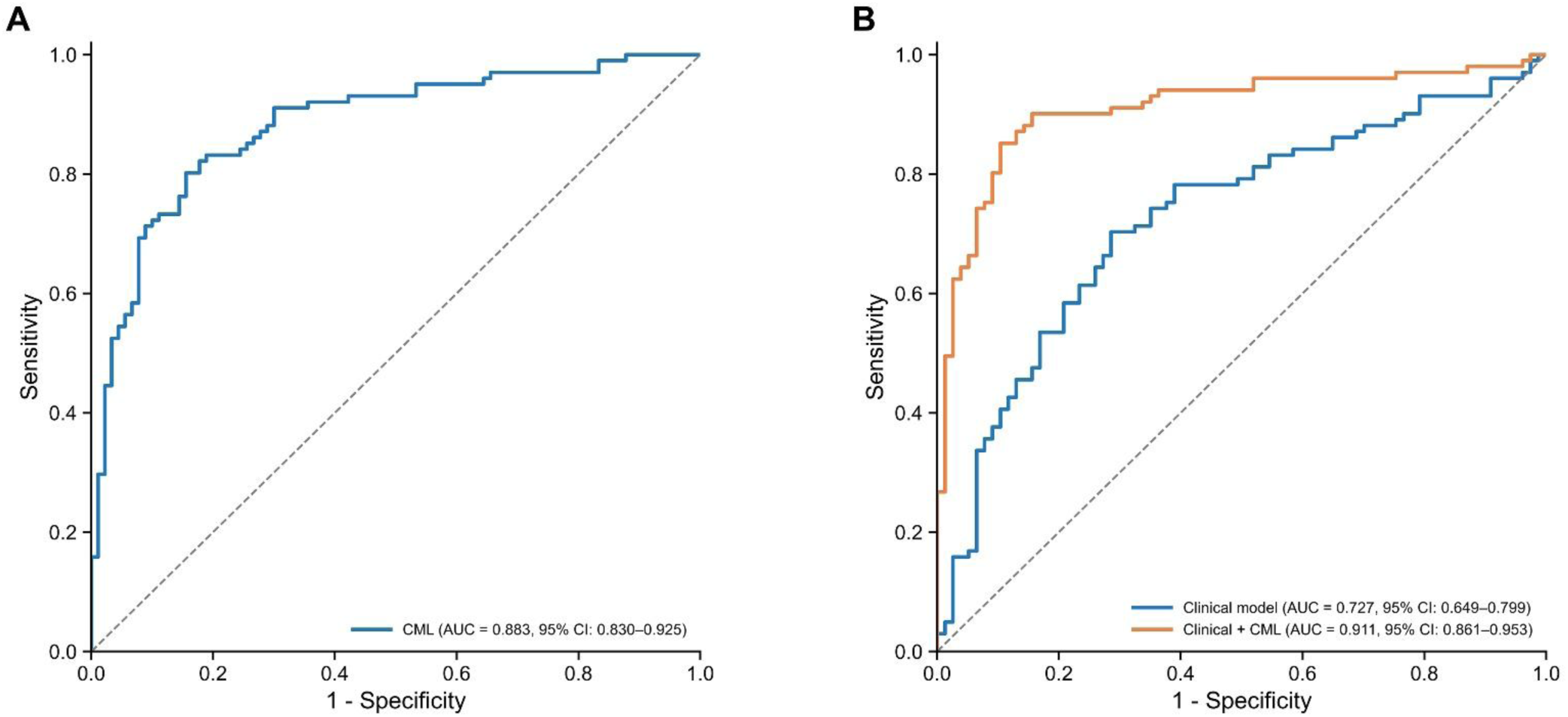
Discriminative performance of serum CML for AD. (A) Receiver operating characteristic (ROC) curve for serum CML alone in 191 participants (101 patients with AD and 90 HC). The area under the curve (AUC) was 0.883 (95% CI, 0.830–0.925). (B) ROC curves comparing a clinical model incorporating age, sex, eGFR, and HbA1c with a combined model additionally including serum CML. The analysis included 178 participants (101 AD and 77 HC) with complete data. ROC curves were generated from out-of-fold predictions using stratified five-fold cross-validation. The AUC was 0.727 (95% CI, 0.649–0.799) for the clinical model and 0.911 (95% CI, 0.861– 0.953) for the combined model. The diagonal dashed lines indicate chance-level discrimination.

### 1.5 Canonical AD-related biomarkers in the single-molecule subcohort

To place the CML finding in the context of established AD-related biological pathways, we next characterized amyloid-, tau-, astroglial-, and neuroaxonal-related serum biomarkers in the randomly selected single-molecule immunoassay subcohort of 31 AD participants and 29 HC. Aβ42 was lower in AD (P=0.000445), whereas Aβ40 was unchanged (P=0.9764), resulting in a markedly lower Aβ42/Aβ40 ratio in AD (P<0.0001) (Figure 5A–C; Supplementary Table 1). Consistent with a tau-related biomarker abnormality, p-tau217 and p-tau181 were both higher in AD (P<0.0001 and P=0.0003068, respectively; Figure 5D, E; Supplementary Table 1). NfL and GFAP were also higher in AD (both P<0.0001; Figure 5F, G; Supplementary Table 1).

**Figure 5.**
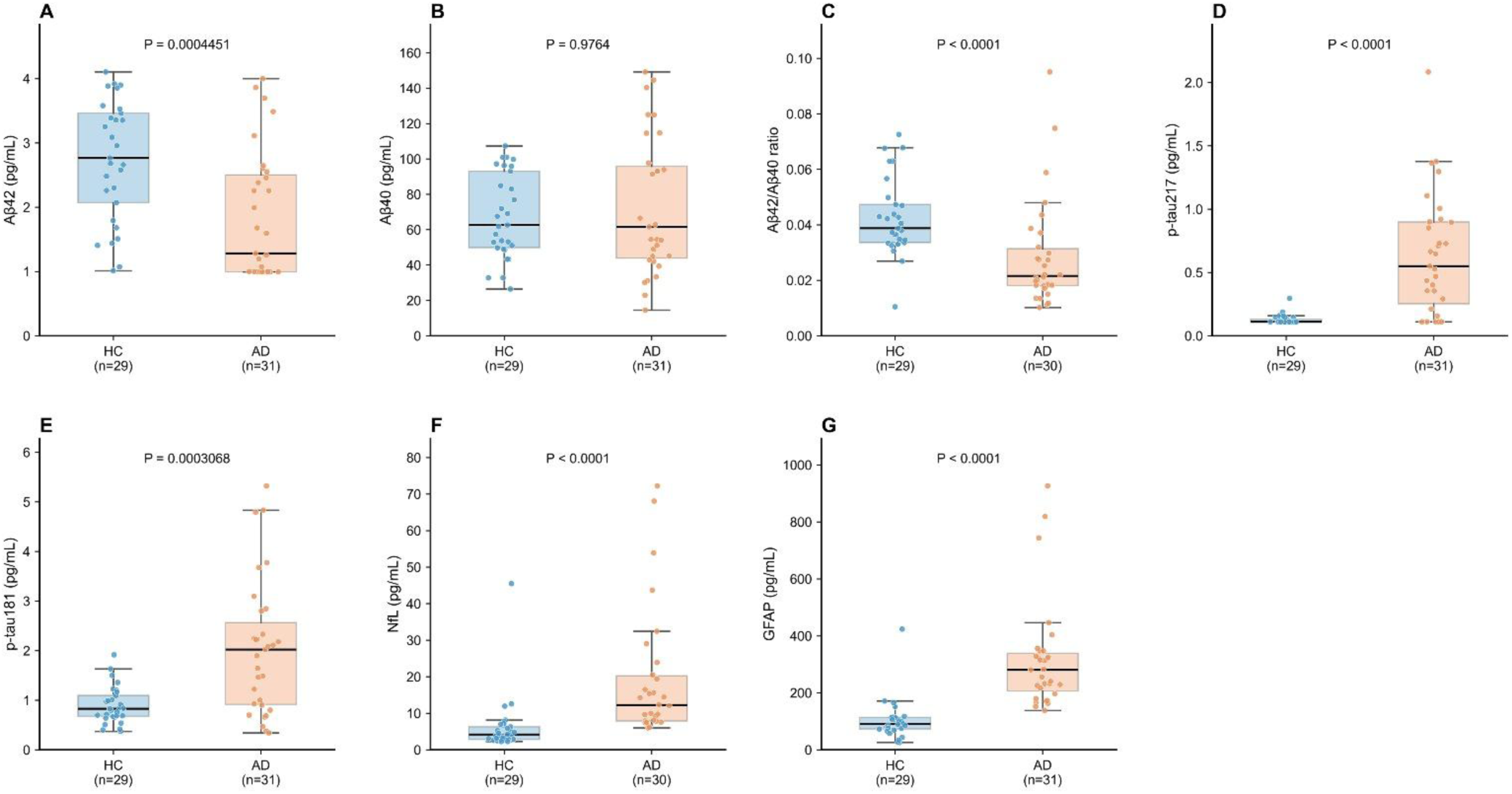
Canonical AD-related biomarkers in the single-molecule subcohort. Serum levels of (A) amyloid-β42 (Aβ42), (B) amyloid-β40 (Aβ40), (C) the Aβ42/Aβ40 ratio, (D) phosphorylated tau217 (p-tau217), (E) phosphorylated tau181 (p-tau181), (F) neurofilament light chain (NfL), and (G) glial fibrillary acidic protein (GFAP) in patients with AD and HC, measured using an ultrasensitive single-molecule array (Simoa) platform. The subcohort included 31 patients with AD and 29 HC participants. Each dot represents an individual participant. Boxes indicate the interquartile range (IQR), with the central horizontal line representing the median. Whiskers extend to the most extreme observations within 1.5×IQR. P values are shown above each panel.

Analyte-specific sample sizes are indicated beneath each panel and in Supplementary Table 1. The Aβ42/Aβ40 ratio and NfL analyses each included 30 AD observations, whereas Aβ42, Aβ40, p-tau217, p-tau181, GFAP, and CML included 31 AD observations; all corresponding biomarker analyses included 29 healthy controls. Collectively, these measurements demonstrate that the subcohort exhibited the expected group-level differences across canonical amyloid- and tau-related biomarkers, together with supportive changes in markers of neuroaxonal injury and astroglial activation, thereby providing biological context for evaluating CML alongside established AD-related signals.

### 1.6 Exploratory ROC comparison in the biomarker subcohort

Finally, to explore whether the systemic glycoxidative information represented by CML might provide discriminatory information complementary to canonical AD-related biomarkers, we evaluated CML in combination with an amyloid-related marker and, separately, a tau-related marker (Figure 6). In the complete CML and Aβ42/Aβ40 dataset (n=59; 30 AD and 29 HC), the cross-validated AUCs were 0.864 for CML, 0.794 for Aβ42/Aβ40, and 0.883 for the combined CML + Aβ42/Aβ40 model (Figure 6A). We next performed the corresponding analysis using p-tau217 as the tau-related comparator. In the complete CML and p-tau217 dataset (n=60; 31 AD and 29 HC), the cross-validated AUCs were 0.878 for CML, 0.844 for p-tau217, and 0.966 for the combined CML + p-tau217 model (Figure 6B). Model performance in both analyses was evaluated using stratified five-fold cross-validation. The slight difference between the two CML AUCs reflects the different complete-case analysis sets. Thus, CML showed only a modest increase in discrimination when combined with Aβ42/Aβ40 but a larger increase when combined with p-tau217. Given the limited subcohort size, however, these combined-model results should be considered exploratory and require independent validation.

**Figure 6.**
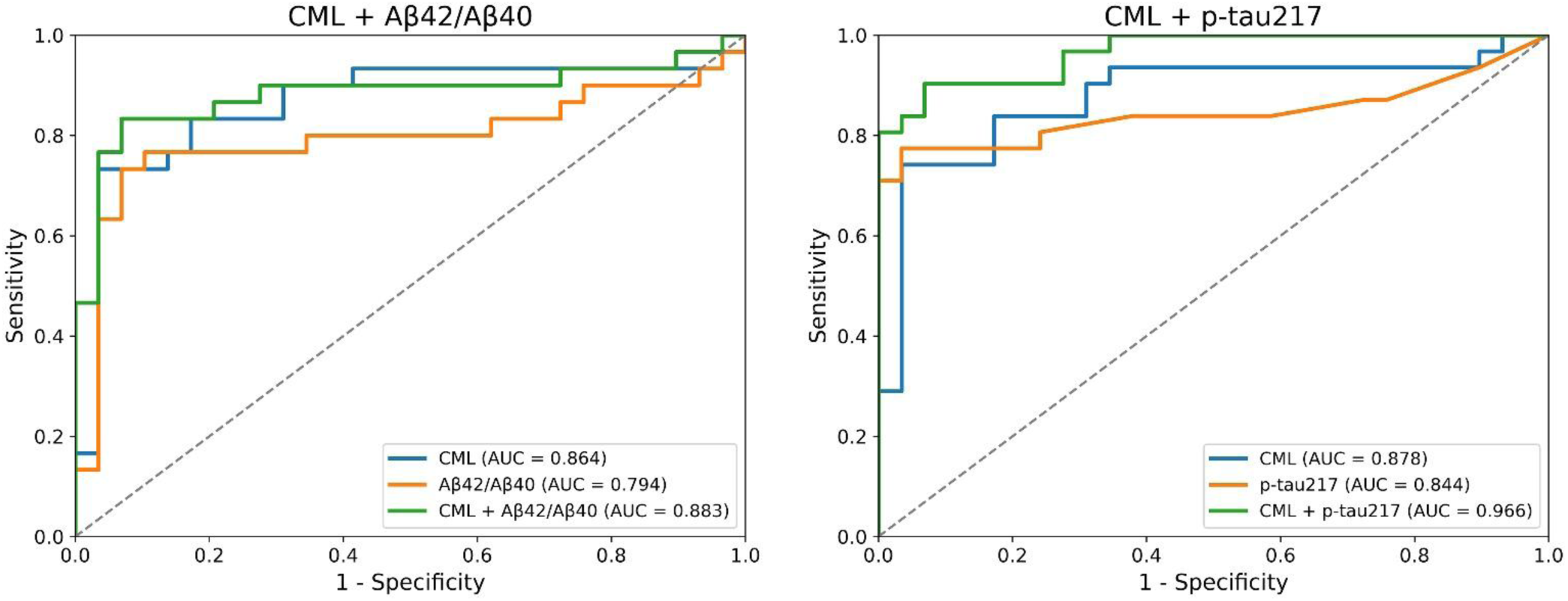
Exploratory ROC analysis of CML combined with established AD-related serum biomarkers. ROC curves comparing the discriminative performance of serum CML, established AD-related biomarkers, and their combinations for distinguishing patients with AD from HC. (A) ROC curves for CML, the Aβ42/Aβ40 ratio, and the combined CML + Aβ42/Aβ40 model (n=59; 30 AD and 29 HC). (B) ROC curves for CML, p-tau217, and the combined CML + p-tau217 model (n=60; 31 AD and 29 HC). Model performance was evaluated using stratified five-fold cross-validation. The area under the ROC curve (AUC) for each model is indicated in the corresponding legend. The dashed diagonal line represents chance-level discrimination.

## Discussion

In this case–control biomarker study, serum CML was markedly lower in patients with clinically diagnosed AD than in healthy controls. The association remained strong after adjustment for age and sex and after further accounting for eGFR and major metabolic covariates, including fasting glucose, HbA1c, BMI, hypertension, and hyperlipidemia. CML alone showed good discrimination (AUC of 0.883). More importantly, in the 178-participant complete-case analysis, stratified five-fold cross-validation yielded an AUC of 0.727 for the clinical model and 0.911 after CML was added. Thus, the lower serum CML signal was not readily explained by the measured demographic, renal, or metabolic differences and provided substantial additional discriminatory information beyond the selected clinical variables. Nevertheless, these results arise from a case–control cohort and still require independent external validation before they can be interpreted as generalizable clinical performance or used to define a diagnostic threshold.

The randomly selected single-molecule immunoassay subcohort showed lower Aβ42 and Aβ42/Aβ40, unchanged Aβ40, and higher p-tau217, p-tau181, glial fibrillary acidic protein (GFAP), and neurofilament light chain (NfL) in AD. These patterns align with established amyloid-, tau-, glial-, and neuroaxonal-associated biomarkers [3–6, 24, 25]. Unlike amyloid and phosphorylated tau markers, which more directly reflect defining AD pathology, CML may capture systemic glycoxidative, carbonyl-related, and metabolic processes. Its potential value is therefore complementary rather than substitutive.

Reduced circulating CML appears paradoxical given reports of increased CML deposition in AD brain tissue. Neuropathological studies reveal heterogeneous cerebral CML distribution across neuronal and glial populations and anatomical regions [14–17]. Immunoreactivity is prominent in cortex and hippocampus, including CA3/CA4, and in astroglial-associated extraneuronal deposits. Intracellular deposits may accompany lipofuscin or vacuolar structures [14–16]. These distinct cellular and molecular pools may differ in formation, retention, and turnover. Thus, regional cerebral accumulation need not parallel serum CML concentrations. This distinction is central to interpreting the apparent discrepancy: immunohistochemical tissue deposition reflects localized modifications, whereas a serum assay measures an assay-dependent circulating fraction at a particular time.

CML deposition also differs from conventional AD pathology. Neurons with prominent neurofibrillary tangles may exhibit little CML, whereas CML-positive neurons can lack substantial tangles. Regional deposition has been inversely associated with Braak stage [15, 17]. Site-specific modification of Aβ42 at Lys16 or Lys28 slows fibrillar aggregation while increasing cytotoxicity, particularly at Lys28 [19]. This may reflect persistent toxic oligomeric intermediates rather than increased reactive oxygen species production. CML’s consequences therefore depend on the substrate, modification site, aggregation state, and cellular environment.

CML may also reflect oxidative lipid injury. Its cerebral co-localization with malondialdehyde links glycoxidative modification to lipid peroxidation [18]. Reactive carbonyls arising from lipid oxidation, alongside carbohydrate-derived pathways, can contribute to CML formation. Local oxidative processes could therefore increase cerebral protein modification without raising circulating CML. The relative contributions of lipid-derived carbonyl chemistry, glycoxidation, and protein turnover in AD remain unresolved. These pathways are not mutually exclusive, and their relative importance may differ across cell types and disease stages.

Glial biology and peripheral trafficking may influence CML homeostasis. Astrocytes and microglia can accumulate CML-modified proteins through local formation, uptake, altered proteostasis, or inflammation [15, 16]. Mossad and colleagues described a gut–blood–brain pathway in which age-associated intestinal barrier changes increased CML exposure and CML impaired microglial mitochondrial function [20]. These findings support intercompartmental interactions but do not establish that cerebral uptake explains our observed serum decrease. Local glycoxidative and lipid-peroxidative formation offers another possibility [17, 18]. Microglial mitochondrial dysfunction is biologically relevant to AD, but the experimental gut–brain findings should not be extrapolated directly to human serum concentrations.

Prior clinical studies underscore the importance of specimen type and molecular fraction. Increased cerebrospinal-fluid CML without a corresponding serum increase suggests partly independent central and peripheral regulation [21]. Increased CML in albumin-free serum proteins during early AD, reduced extracellular-vesicle-associated CML in moderate disease [22], and increased plasma CML with sex-related variation [23] have all been reported. Serum versus plasma, free versus protein-bound CML, extracellular-vesicle-associated CML, disease stage, analytical platform, renal function, metabolic status, and cohort composition may all contribute to heterogeneity. CML should not be treated as a homogeneous analyte across biological compartments and methods.

Our assay comparisons further support this distinction. The in-house competitive enzyme-linked immunosorbent assay (ELISA) responded concentration-dependently to purified free CML and CML-modified bovine serum albumin, whereas the commercial assay showed little response to CML-modified protein under tested conditions (Supplementary Figure 3). The in-house method therefore recognizes a broader immunoreactive pool, and the commercial method appears predominantly sensitive to a free-CML-associated fraction. However, absent response to CML-BSA does not prove absolute specificity for free CML across all modified proteins. Both assays yielded lower AD signals and a moderate positive rank correlation in 60 paired samples (Supplementary Figure 2). Their differing calibrations and detectable pools preclude direct comparison of absolute concentrations or units. Our primary finding concerns the commercial assay-derived signal. Thus, concordant between-group direction and rank-order correlation should be distinguished from analytical agreement, and neither immunoassay alone establishes absolute total CML content.

Several nonexclusive mechanisms could explain lower serum CML. CML-related modifications occur in circulating proteins, blood cells, extracellular vesicles, peripheral tissues, and brain [15, 22, 26–28]. Changes in protein turnover, tissue retention, uptake, or vesicle trafficking might redistribute immunoreactive CML. Diet, intestinal absorption, inflammation, systemic glycoxidative flux, and clearance could also alter circulating concentrations [8–13, 20]. Lower serum CML need not imply lower whole-body formation or cerebral burden. These remain hypotheses because we did not quantify CML flux, identify modified proteins, or sample matched central and peripheral compartments.

Renal and metabolic confounding warrant attention. Impaired renal elimination generally increases circulating CML [12, 13], yet eGFR was modestly lower in AD despite markedly lower serum CML. Within-group CML–eGFR correlations were nonsignificant (AD: ρ=0.012, P=0.904; controls: ρ=0.202, P=0.060; Supplementary Figure 1). Adjustment for eGFR, glycemic indices, and metabolic variables did not materially weaken the association. Nevertheless, diet, medications, nutritional status, smoking, inflammation, and intestinal physiology could contribute residual confounding. These findings reduce the plausibility of a simple renal explanation but do not exclude more complex interactions among renal function, nutrition, and inflammation.

A compartment-dependent model may reconcile localized cerebral deposition with reduced circulating CML without assuming direct serum-to-brain sequestration. Regional formation, protein-specific modification, cellular uptake, vesicular trafficking, and systemic clearance may change independently. Testing this model requires orthogonal quantification of free and protein-bound CML in serum or plasma, blood cells, extracellular vesicles, cerebrospinal fluid, and, where possible, brain tissue. Targeted mass spectrometry, identification of CML-modified proteins, and longitudinal sampling could distinguish formation from redistribution and clearance. Paired measurements would be especially informative if accompanied by detailed information on protein modification, sample handling, and renal function.

In our subcohort, the expected abnormalities in Aβ42/Aβ40 and p-tau217 confirmed canonical amyloid- and tau-related signals [24], supporting the idea that CML captures a biological dimension not identical to amyloid deposition, tau phosphorylation, astroglial activation, or neuroaxonal injury. Exploratory combined-marker analyses suggested potential complementarity. CML plus Aβ42/Aβ40 yielded an AUC of 0.883 versus 0.864 for CML alone in 59 participants. CML plus p-tau217 yielded an AUC of 0.966 versus 0.878 for CML and 0.844 for p-tau217 individually in 60 participants.

Despite five-fold cross-validation, these small analyses are vulnerable to sampling variability and model optimism. Incremental value beyond validated p-tau217 and Aβ42/Aβ40 assays requires independent evaluation in cognitively unimpaired individuals, mild cognitive impairment, AD, and non-AD dementias. The apparent improvement should not be regarded as a validated diagnostic gain until its magnitude and uncertainty are reproduced prospectively.

The strengths of this study include a primary cohort approaching 200 participants, adjustment for major measured CML determinants, a randomly selected biomarker subcohort, cross-validated discrimination analyses, and experimental characterization of assay recognition. The limitations include the single-cohort, cross-sectional case–control design, which precludes causal inference and longitudinal prediction, as well as absent external validation, residual confounding, and small exploratory analyses. Neither ELISA fully resolves circulating CML molecular pools or identifies modified proteins. The full cohort also lacked uniform amyloid PET, tau PET, or cerebrospinal-fluid confirmation, limiting generalization to biomarker-defined AD. In addition, clinical diagnosis may introduce classification error relative to biological AD criteria, and the direction of any resulting bias is uncertain.

Future studies should validate the association across cohorts and analytical platforms, determine disease specificity, and follow CML trajectories across cognitive aging, mild cognitive impairment, and AD. Integrating molecularly resolved CML measurements with lipidomics, metabolomics, renal and dietary measures, intestinal-barrier biology, and microbiome profiles could clarify its biological origins. Prospective studies should test incremental information beyond established blood biomarkers and associations with progression or treatment response. Such studies should prespecify analytical targets and normalization procedures to avoid conflating free CML with protein-bound immunoreactivity.

In conclusion, a lower free-CML-sensitive serum signal was independently associated with clinically diagnosed AD and showed exploratory complementarity with established biomarker patterns. These findings motivate investigation of compartment-dependent CML homeostasis, but mechanisms, disease specificity, and clinical utility require longitudinal, biomarker-defined, externally validated studies.

## Acknowledgement

This work was supported by fundings from the Yongjiang Talent Introduction Programme (Young Innovative Talent Project, 2021A-012-G), Zhejiang Provincial Natural Science Foundation (LQ24H160003), the Medical Scientific Research Foundation of Zhejiang Province (2024KY351), the Ningbo Natural Science Foundation (2023J365), and the Bei’An Talent Programme of Jiangbei District, Ningbo (2023RC004).

## Data Availability Statement

The de-identified data supporting the findings of this study are available from the corresponding author upon reasonable request, subject to applicable ethical and institutional requirements.

## Supporting Information

**Supplementary Table 1.**
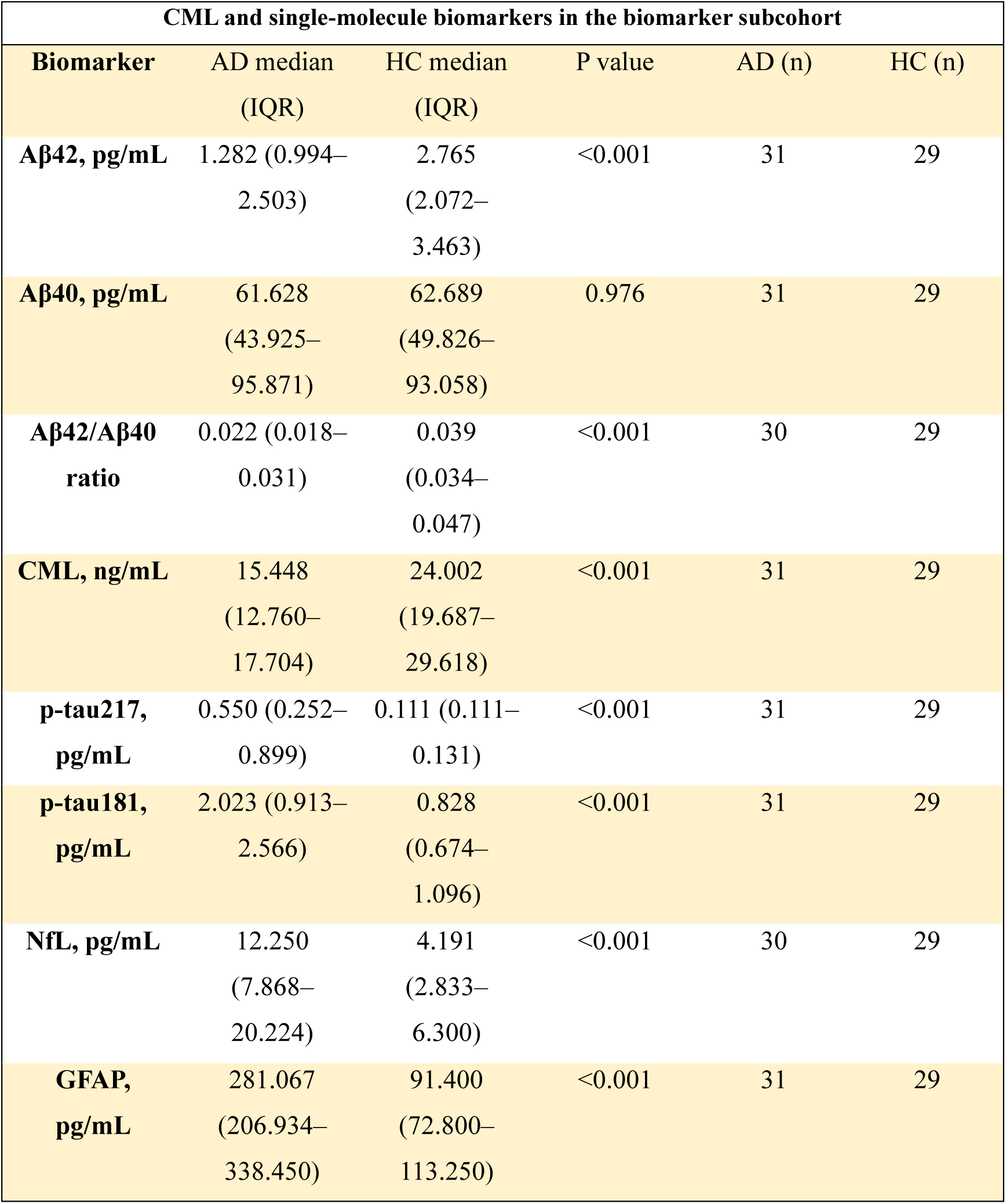
CML and single-molecule biomarkers in the biomarker subcohort. Data are presented as median (interquartile range, IQR). Differences between the AD and HC groups were assessed using the two-sided Mann–Whitney U tests. The table includes CML, Aβ42, Aβ40, Aβ42/Aβ40, p-tau217, p-tau181, NfL, and GFAP. Analyte-specific sample sizes are shown because the final analysis sets differed slightly across biomarkers. Results reported below the assay lower limit of quantification were handled as described in the Methods. Vendor-supplied reference intervals that were not validated for serum were not used to classify participants as biomarker-positive or biomarker-negative. AD, Alzheimer’s disease; HC, healthy control; CML, N^ε^-(carboxymethyl)lysine; NfL, neurofilament light chain; GFAP, glial fibrillary acidic protein.

| **CML and single-molecule biomarkers in the biomarker subcohort** |  |  |  |  |  |
| **Biomarker** | AD median (IQR) | HC median (IQR) | P value | AD (n) | HC (n) |
|---|---|---|---|---|---|
| **Aβ42, pg/mL** | 1.282 (0.994–2.503) | 2.765 (2.072–3.463) | <0.001 | 31 | 29 |
| **Aβ40, pg/mL** | 61.628 (43.925–95.871) | 62.689 (49.826–93.058) | 0.976 | 31 | 29 |
| **Aβ42/Aβ40 ratio** | 0.022 (0.018–0.031) | 0.039 (0.034–0.047) | <0.001 | 30 | 29 |
| **CML, ng/mL** | 15.448 (12.760–17.704) | 24.002 (19.687–29.618) | <0.001 | 31 | 29 |
| **p-tau217, pg/mL** | 0.550 (0.252–0.899) | 0.111 (0.111–0.131) | <0.001 | 31 | 29 |
| **p-tau181, pg/mL** | 2.023 (0.913–2.566) | 0.828 (0.674–1.096) | <0.001 | 31 | 29 |
| **NfL, pg/mL** | 12.250 (7.868–20.224) | 4.191 (2.833–6.300) | <0.001 | 30 | 29 |
| **GFAP, pg/mL** | 281.067 (206.934–338.450) | 91.400 (72.800–113.250) | <0.001 | 31 | 29 |

**Supplementary Figure 1.**
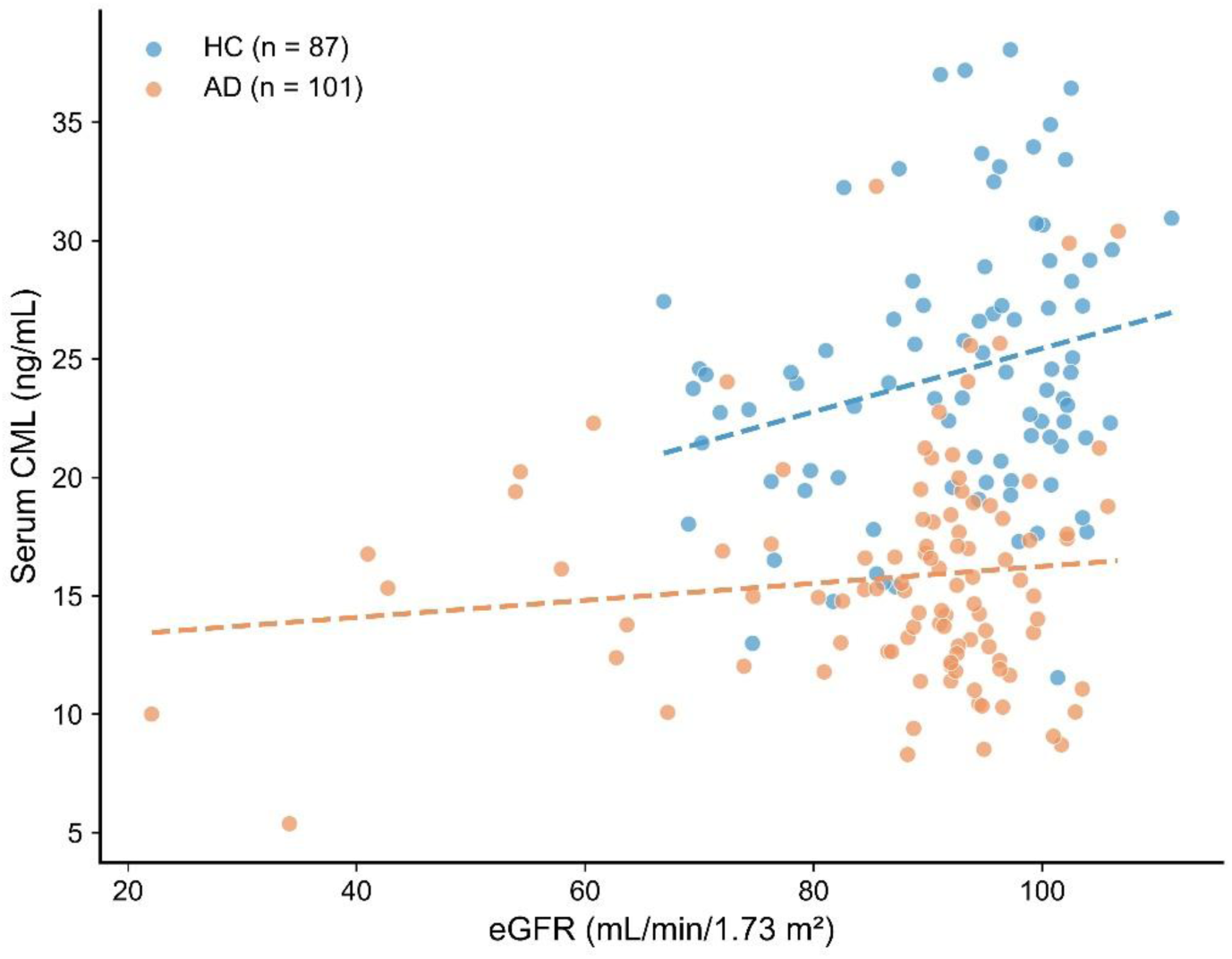
Serum CML in relation to renal function. Scatter plot showing the relationship between serum N^ε^-(carboxymethyl)lysine (CML) concentrations and estimated glomerular filtration rate (eGFR) in patients with Alzheimer’s disease (AD; n=101) and healthy controls (HC; n=87). Each point represents an individual participant. Blue and orange symbols indicate HC and AD participants, respectively. Dashed lines represent group-specific linear regression trends. Spearman rank correlation analysis was performed separately for each group. No significant correlation was observed in AD (ρ=0.012, P=0.904) or HC (ρ=0.202, P=0.060).

**Supplementary Figure 2.**
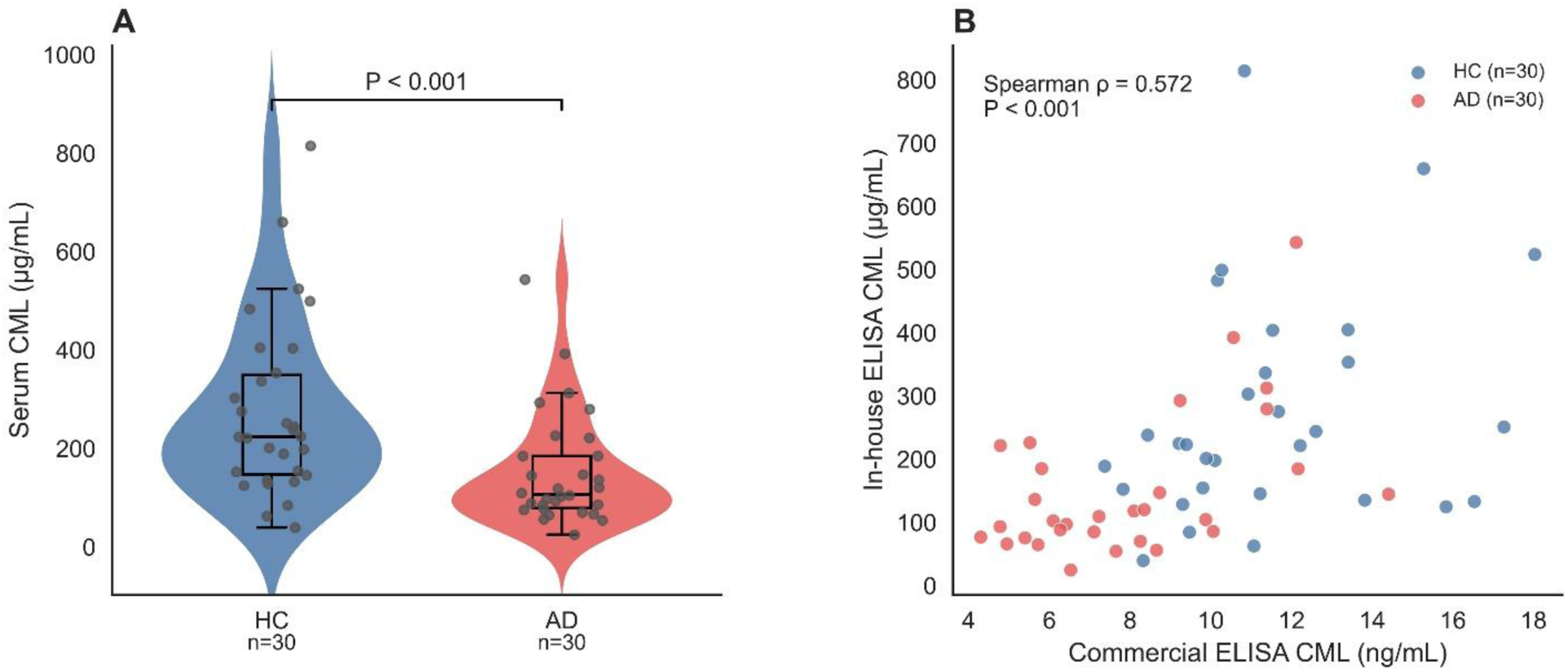
Comparison of serum CML measurements using an in-house competitive ELISA and a commercial ELISA kit. (A) Serum CML concentrations measured using the in-house competitive ELISA in healthy controls (HC, n=30) and patients with Alzheimer’s disease (AD, n=30). Violin plots represent the estimated data distributions, box plots indicate the median and interquartile range, and individual dots represent individual serum samples. Statistical significance was assessed using a two-sided Mann–Whitney U test. (B) Correlation between serum CML concentrations measured using the commercial ELISA kit and the in-house competitive ELISA in the same 60 serum samples. Each dot represents an individual serum sample, with colors indicating the HC and AD groups. Spearman rank correlation analysis revealed a moderate positive correlation between the two assays (ρ=0.572, P<0.001). The absolute concentration scales and reported units differ because the assays have different molecular target profiles and calibration systems: analytical experiments showed that the in-house assay detects both free CML and CML-modified protein, whereas the commercial kit showed little detectable response to CML-BSA (Supplementary Figure S3). Accordingly, the paired analysis evaluates concordance in relative CML variation rather than direct numerical equivalence between assays.

**Supplementary Figure 3.**
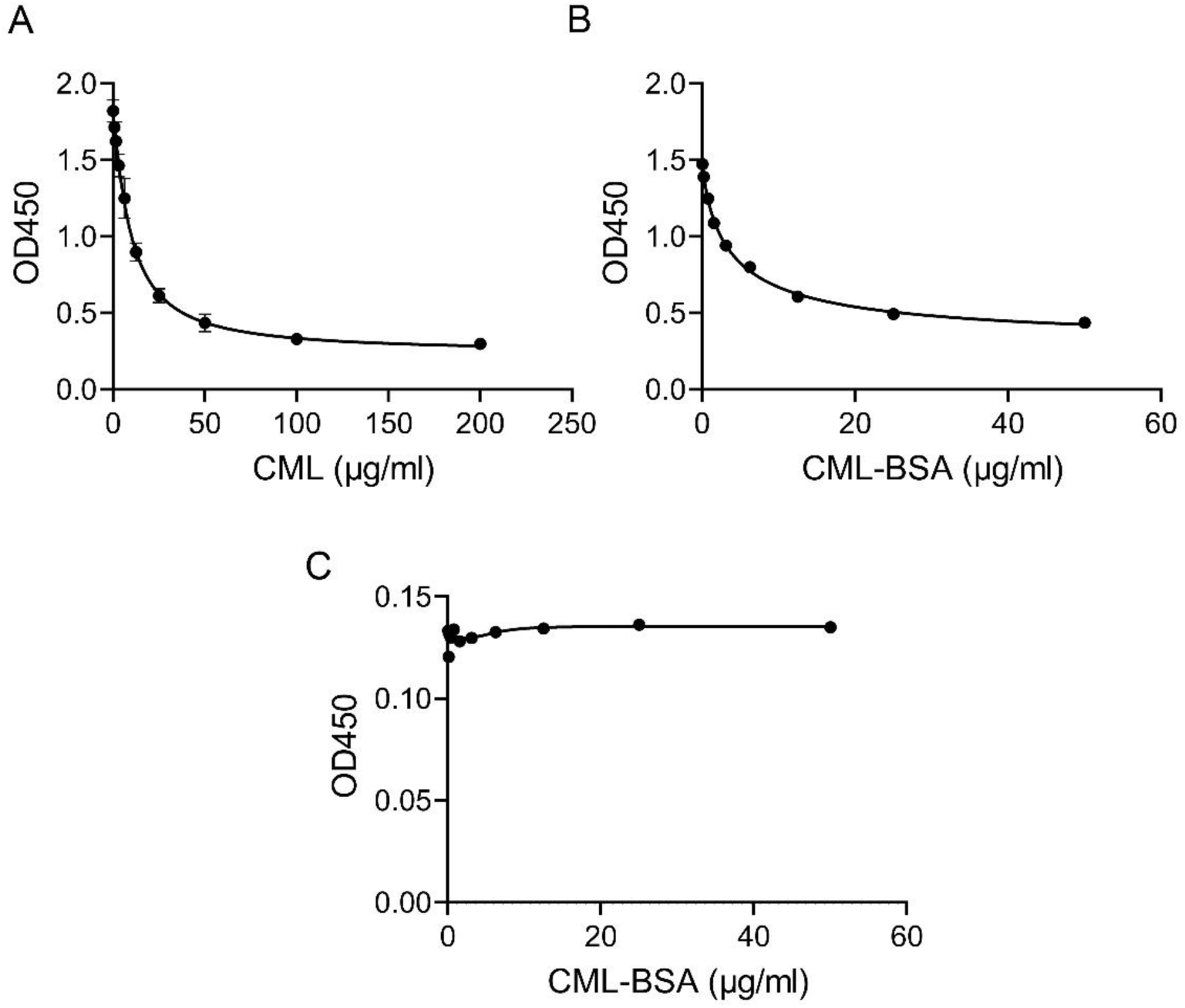
Differential recognition of free and protein-bound CML by the in-house assay and commercial CML kit. Analytical response curves were generated using purified free Nε-(carboxymethyl)lysine (CML) or CML-modified bovine serum albumin (CML-BSA). (A) Concentration-response curve for free CML measured using the in-house indirect competitive ELISA. Increasing free CML concentrations produced a progressive decrease in OD450, demonstrating detection of free CML. (B) Concentration-response curve for CML-BSA measured using the in-house assay. Increasing CML-BSA concentrations likewise produced a concentration-dependent decrease in OD450, demonstrating recognition of protein-bound CML epitopes. Together, panels A and B show that the in-house assay responds to both free CML and CML-modified protein. (C) CML-BSA tested using the commercial CML ELISA kit. OD450 remained essentially unchanged across the tested CML-BSA concentration range, indicating little detectable response to protein-bound CML under these assay conditions. These analytical findings indicate that the in-house assay measures a broader immunoreactive CML pool comprising free and protein-bound CML, whereas the commercial kit predominantly reflects a free-CML-sensitive pool. This difference in molecular target profile and calibration accounts for the markedly different absolute concentration scales observed between the two methods in Supplementary Figure S2; values from the two assays should therefore not be interpreted as directly interchangeable.

